# Physical violence during pregnancy in Peru: Proportion, geographical distribution, and associated factors

**DOI:** 10.1101/2020.05.27.20115030

**Authors:** Mercedes Joselyn Nuñez-Ochoa, Victor Hugo Moquillaza-Alcantara, Clara Margarita Diaz Tinoco

## Abstract

**Objective:** Estimate the proportion, geographic distribution and sociodemographic factors associated with physical violence during pregnancy between 2016 and 2018.

**Material and methods:** Secondary analysis of the Demographic and Family Health Survey, which included respondents whether they presented physical violence during pregnancy in the last 12 months.

**Results:** The proportion of physical violence was 9,9% [95%CI:9,6–10,4%] during 2016, 9,2% [95%CI:8,8–9,6%] during 2017 and 8,6% [95%CI:8,3–8,9%] during 2018, The regions with the highest proportion were Puno, Arequipa and Apurímac during the 3 years. Among the associated factors, the residue in rural areas (RP:0,49; p=0,011) and be “very rich” (RP:0,63; p=0,029) was protective; while they were at risk of not presenting studies (RP:1,87; p=0,014), the cohabiting marital status (RP:1,51; p=0,001), separated (RP:3,56; p<0,001), showing an age between 40 a 49 years (RP:1,79; p=0,012) and that partner drinks alcohol (RP:1,61; p<0,001).

**Conclusion:** The proportion of violence in Peru has been decreasing. The factors that predispose this phenomenon are the wealth index, educational level, marital status, and the age of the pregnant woman.

## Introduction

Violence during pregnancy is a global health problem that usually occurs in poverty settings, resulting in not only physical damage, but also psychiatric morbidity, with a higher proportion of disorders such as post-traumatic stress and postpartum depression^1–3^. The proportion of this type of violence differs between countries, reaching 30% in European countries^4–7^ and 36% in African countries^8^. In Peru, few studies have focused on aggression during pregnancy, among them some texts mention that in the hospitals of the cities of Moquegua^9^ and Ica^10^ the proportion reaches 9.5% and 28%, respectively; however, there is no report that estimates the values at the national level.

Due to the period in which violence occurs, there are related obstetric repercussions, such as the risk of preterm delivery, low birth weight, small for gestational age and insufficient prenatal controls^11^. These results are precisely those that can be avoided with timely and specific interventions in those high-risk populations, which according to previous studies is not yet effective^12^. Therefore, the objective of this study is to identify the proportion and geographical distribution of cases of violence during pregnancy in Peru and its associated sociodemographic factors during the period 2016 to 2018.

## Material and methods

Secondary-based analysis of the Demographic and Family Health Survey (ENDES) of Peru for the last 3 years: 2016, 2017 and 2018; which are freely available on the website of the National Institute of Statistics and Informatics (http://iinei.inei.gob.pe/microdatos/). The databases of module 64 “Characteristics of the home” (RECH0 and RECH1), module 66 “Basic data of MEF” (REC0111) and module 73 “Family Violence” (REC84DV) of the years were considered for analysis.

Those women between the ages of 15 and 49 who have ever been pregnant entered the study and answered the question “Has anyone ever physically hit, slapped, kicked or mistreated you while pregnant?” (question 1019 of the questionnaire, variable D118Y). The sample, as specified in the technical sheet, was probabilistic of the stratified, three-stage and self-weighted type.

The proportion of “physical violence during pregnancy” was evaluated using the affirmative or negative answer to question 1019 of the family violence survey. Likewise, it was also evaluated whether the participant sought help based on her answer to the question “When they have mistreated you, have you asked for help from people close to you?” (question 1022 of the questionnaire, variable D119Y). The following were included as factors: the region of origin (variable HV023), maximum level of education approved (HV106), area of residence (HV025), place of residence (HV026), marital status (HV115), wealth index (V190), alcohol consumption by the couple (D113) and age (HV105), the latter being categorized in periods of 10 years. All variables were available in the 3 years evaluated.

The analysis was carried out in the STATA version 14 software, where the stratification, clusters and weighting factors established by ENDES were taken into account, in order to estimate all the results according to complex sampling and considering the svy command for analysis. Base binding was performed considering the binding variables HHID, HVIDX and CASEID. Proportions with their respective confidence intervals were estimated for each of the years evaluated; Likewise, when analysing the proportion of violence by region, the confidence intervals and the coefficient of variation (CV) were considered, assuming that CV> 15 as “referential”.

Associated factors were evaluated using a generalized linear model of the Poisson family for complex samples with a link (log), reporting the sense of association using the adjusted Prevalence Ratio (aRP) and its respective 95% confidence interval. The geographical distribution was evaluated using the QGIS software, where the geographic reference of the National Institute of Statistics and Informatics was taken and cut points for the proportions were established, which were generated from the maximum and minimum values found in the 3 years. of studies.

The analysed bases are freely accessible, and the records did not present identifiable information of the participants, for which the approval of an ethics committee was not required. Since the present investigation derives from an undergraduate thesis,^13^ it was reviewed and approved prior to its execution by the research committee of the obstetrics school of the Universidad Nacional Mayor de San Marcos, as part of the thesis evaluation process. university.

## Results

The number of participants who met the selection criteria were 21,736, 22,130, and 23,315 women during 2016, 2017, and 2018, respectively. During 2016, of all those participants who presented a pregnancy in the last 12 months, there was a proportion of 9.9% [95%CI: 9.6–10.4%] who reported having presented physical violence; value that reached 9.2% [95%CI: 8.8–9.6%] and 8.6% [95%CI: 8.3–8.9] during the years 2017 and 2018, respectively. Likewise, the proportion of participants that when perceiving physical violence never asked for help was evaluated, which presented a maximum value during 2017, of 45.5% [95%CI: 43.4–47.7%], and the lowest value of 43.3% [95%CI: 41.2–45.4%] during 2016. (Figure 1).

**Figure 1.**
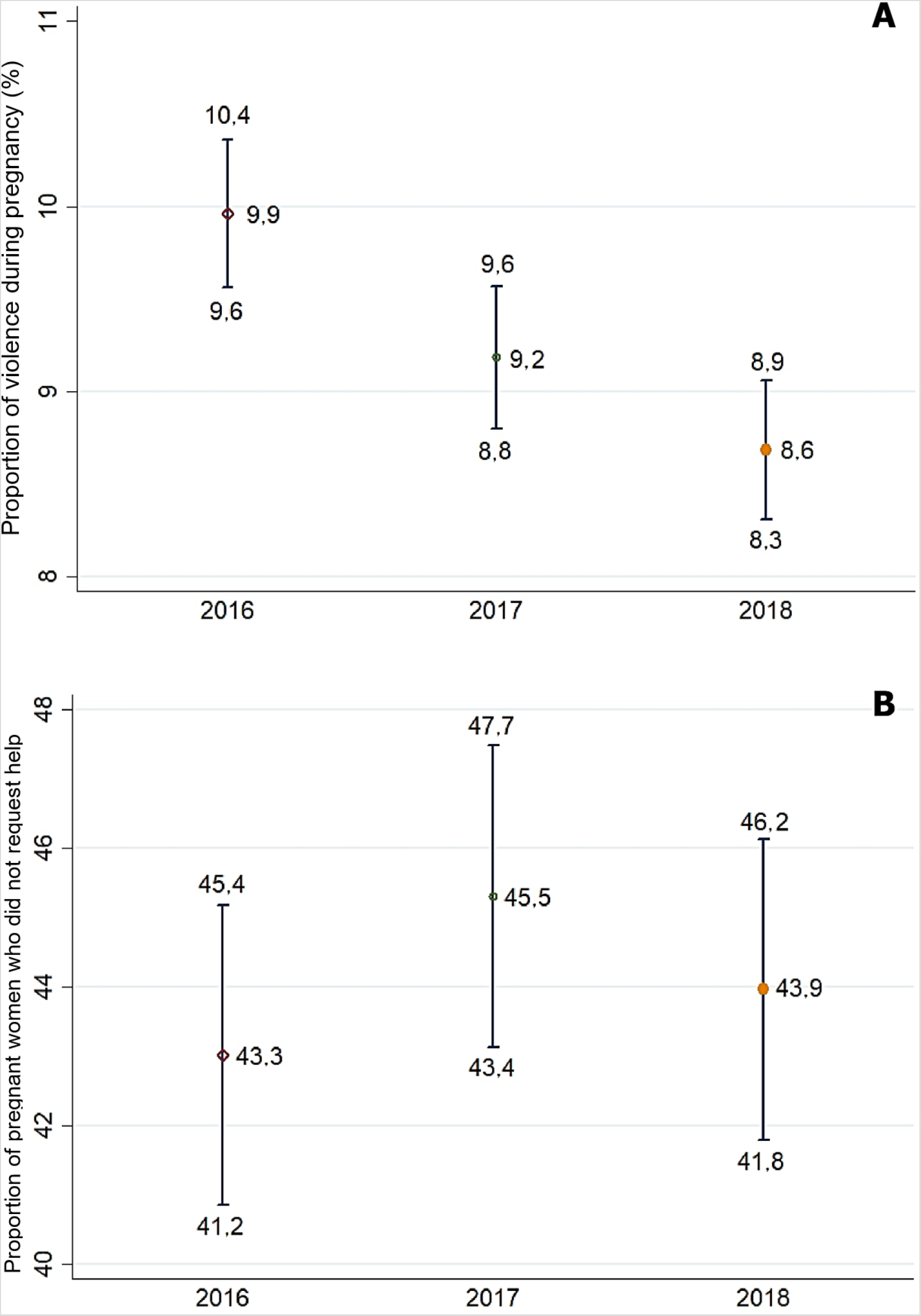
Proportion of A) violence during pregnancy and B) pregnant women victims who did not request help, Peru 2016–2018.

Regarding the proportion of violence during pregnancy in the regions of Peru during the last 3 years, those with high indicators that were reducing their percentages were the regions of Apurímac (from 15.8% to 13.2%), Cusco (from 15.0% to 12.1%) and Tumbes (from 14.2% to 11.9%). On the other hand, the region of La Libertad presented a proportion of violence of 3.5% during 2016, which increased to 7.7% during 2018. Just in this last year those regions with the lowest proportion of violence during the Pregnancy were Cajamarca (5.1%), Tacna (5.1%) and Pasco (5.5%). For its part, the capital, Lima, showed a reduction from 10.3% to 9.2% during the period 2016 to 2018. (Figure 2, Table 1)

**Figure 2.**
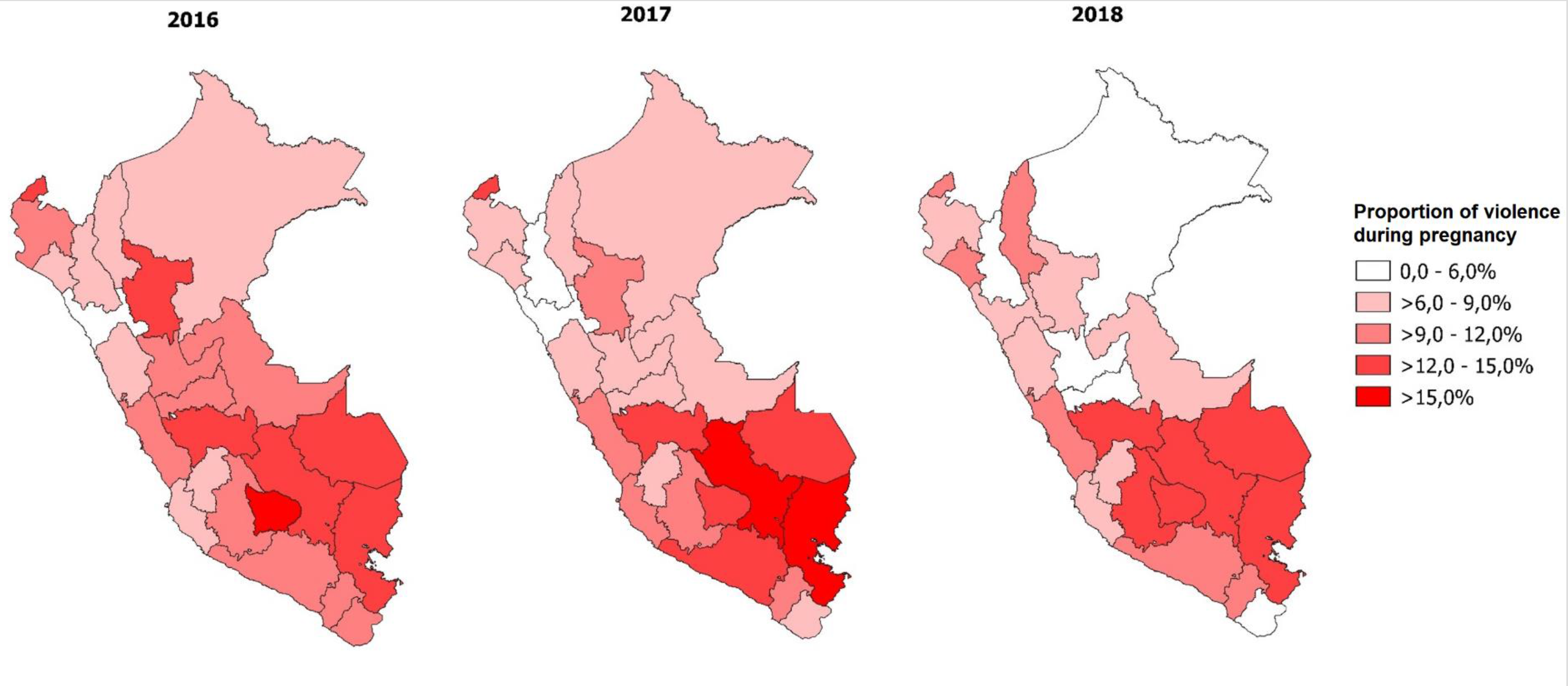
Geographical distribution of violence during pregnancy in the regions of Peru, 2016 – 2018.

**Table 1.** Proportion of violence during pregnancy in the regions of Peru during the period 2016 – 2018.

|  | 2016 |  |  | 2017 |  |  | 2018 |  |  |
| --- | --- | --- | --- | --- | --- | --- | --- | --- | --- |
|  | n<br>(N)* | % **<br>[IC95%] | CV (%) | N<br>(N)* | % **<br>[IC95%] | CV (%) | n<br>(N)* | % **<br>[IC95%] | CV (%) |
| Amazonas | 65<br>(926) | 7,0<br>[5,4-9,1] | 13.0 | 65<br>(907) | 6,8<br>[5,1-8,9] | 14.4 | 83<br>(960) | 9,1<br>[7,0-11,7] | 12.9 |
| Ancash | 57<br>(724) | 7,6<br>[5,6-10,3] | 15.3 | 53<br>(738) | 8,0<br>[5,7-11,3] | 17.3 | 63<br>(846) | 7,9<br>[5,6-11,1] | 17.3 |
| Apurimac | 110<br>(761) | 15,8<br>[12,7-19,5] | 10.8 | 109<br>(785) | 14,5<br>[11,9-17,5] | 9.8 | 105<br>(819) | 13,2<br>[10,4-16,6] | 11.9 |
| Arequipa | 77<br>(730) | 9,8<br>[7,5-12,8] | 13.4 | 79<br>(779) | 12,2<br>[9,3-15,9] | 13.7 | 79<br>(807) | 10,4<br>[7,9-13,5] | 13.2 |
| Ayacucho | 96<br>(905) | 10,7<br>[8,5-13,2] | 11.1 | 82<br>(884) | 9,2<br>[7,1-11,8] | 12.8 | 120<br>(943) | 12,8<br>[10,0-16,2] | 12.1 |
| Cajamarca | 44<br>(730) | 6,1<br>[4,3-8,7] | 17.6 | 49<br>(793) | 5,9<br>[4,2-8,4] | 17.2 | 41<br>(889) | 5,1<br>[3,4-7,5] | 20.1 |
| Callao | 71<br>(805) | 9,9<br>[7,6-12,9] | 13.5 | 78<br>(809) | 11,7<br>[8,6-15,7] | 15.2 | 65<br>(803) | 9,4<br>[6,9-12,7] | 15.5 |
| Cusco | 105<br>(700) | 15,0<br>[12,3-18,3] | 10.1 | 94<br>(682) | 15,3<br>[11,0-20,9] | 16.1 | 95<br>(765) | 12,1<br>[9,3-15,6] | 13.0 |
| Huancavelica | 66<br>(716) | 8,9<br>[6,7-11,6] | 13.8 | 50<br>(671) | 7,8<br>[5,7-10,7] | 15.9 | 62<br>(801) | 8,7<br>[6,5-11,4] | 14.1 |
| Huanuco | 82<br>(874) | 9,9<br>[7,9-12,4] | 11.4 | 75<br>(903) | 8,3<br>[6,5-10,6] | 12.3 | 57<br>(916) | 5,9<br>[4,3-7,9] | 15.5 |
| Ica | 80<br>(889) | 8,7<br>[6,9-10,8] | 11.3 | 77<br>(884) | 9,3<br>[6,8-12,5] | 15.3 | 72<br>(866) | 7,7<br>[5,7-10,4] | 15.4 |
| Junin | 94<br>(807) | 12,3<br>[9,6-15,6] | 12.2 | 80<br>(818) | 12,4<br>[9,3-16,4] | 14.3 | 81<br>(871) | 12,6<br>[9,4-16,7] | 14.6 |
| La libertad | 31<br>(784) | 3,5<br>[2,4-5,1] | 19.1 | 46<br>(837) | 4,9<br>[3,9-7,0] | 18.4 | 60<br>(844) | 7,7<br>[5,7-10,3] | 15.1 |
| Lambayeque | 49<br>(869) | 7,0<br>[4,6-10,5] | 20.7 | 62<br>(861) | 7,8<br>[5,7-10,5] | 15.3 | 62<br>(890) | 9,3<br>[6,7-12,7] | 15.9 |
| Lima | 248<br>(2404) | 10,3<br>[8,5-12,3] | 9.3 | 223<br>(2413) | 9,8<br>[7,9-11,9] | 10.4 | 247<br>(2835) | 9,2<br>[7,7-11,1] | 9.4 |
| Loreto | 65<br>(877) | 8,2<br>[5,8-11,5] | 17.2 | 58<br>(885) | 6,9<br>[4,9-9,7] | 16.7 | 51<br>(882) | 5,7<br>[4,2-7,8] | 15.9 |
| Madre de Dios | 97<br>(773) | 13,2<br>[10,7-16,1] | 10.3 | 111<br>(852) | 13,1<br>[10,3-16,4] | 11.7 | 85<br>(821) | 12,5<br>[9,6-16,1] | 13.0 |
| Moquegua | 77<br>(744) | 10,9<br>[8,3-14,1] | 13.4 | 75<br>(756) | 10,3<br>[7,5-13,9] | 15.6 | 65<br>(780) | 9,6<br>[6,9-13,3] | 16.8 |
| Pasco | 69<br>(779) | 10,0<br>[7,6-13,2] | 13.9 | 66<br>(767) | 8,2<br>[6,0-11,1] | 15.4 | 42<br>(753) | 5,5<br>[3,8-7,9] | 18.6 |
| Piura | 79<br>(881) | 9,3<br>[7,2-12,1] | 13.2 | 66<br>(876) | 7,2<br>[5,3-9,7] | 15.4 | 70<br>(919) | 7,8<br>[5,9-10,3] | 14.2 |
| Puno | 88<br>(616) | 14,4<br>[11,2-18,4] | 12.4 | 101<br>(649) | 16,4<br>[12,7-20,8] | 12.4 | 81<br>(659) | 14,7<br>[11,3-18,8] | 12.8 |
| San Martín | 106<br>(868) | 13,4<br>[10,8-16,5] | 10.6 | 89<br>(927) | 10,7<br>[8,1-14,1] | 14.1 | 89<br>(932) | 8,0<br>[6,1-10,4] | 13.6 |
| Tacna | 92<br>(784) | 11,6<br>[9,1-14,7] | 12.1 | 58<br>(790) | 7,4<br>[5,2-10,4] | 17.3 | 56<br>(851) | 5,1<br>[3,5-7,4] | 19.2 |
| Tumbes | 122<br>(913) | 14,2<br>[11,4-17,5] | 10.8 | 112<br>(932) | 13,4<br>[10,8-16,6] | 10.9 | 110<br>(907) | 11,9<br>[9,3-15,2] | 12.5 |
| Ucayali | 95<br>(877) | 11,4<br>[9,4-13,7] | 9.5 | 73<br>(932) | 8,7<br>[6,3-11,9] | 15.9 | 73<br>(956) | 7,8<br>[5,8-10,5] | 14.9 |

n: Violent during pregnancy; N: Total of women surveyed; 95%CI: 95% confidence interval; CV: Coefficient of variation
\* Absolute frequencies; \*\* Weighted proportions according to the complex sample of ENDES 2018

Finally, Table 2 reports the factors associated with violence during pregnancy. Regarding the highest level of studies approved, the proportion of violence increased when they had a lower academic level, where pregnant women without studies were up to 87% more likely to present violence compared to those with higher education. Also, residing in rural areas reduced the probability by 51%. Another factor with associated categories was marital status, where when taking married status as a reference, being cohabiting, divorced, never married, and separated were risk factors for violence. Regarding age, it was found that those between 40 and 49 years old have a greater probability (79%) of violence. When evaluating the wealth index, it was found that those who are categorized as “very rich” have a 37% less probability of violence, while alcohol consumption increases this probability (61%).

**Table 2.**
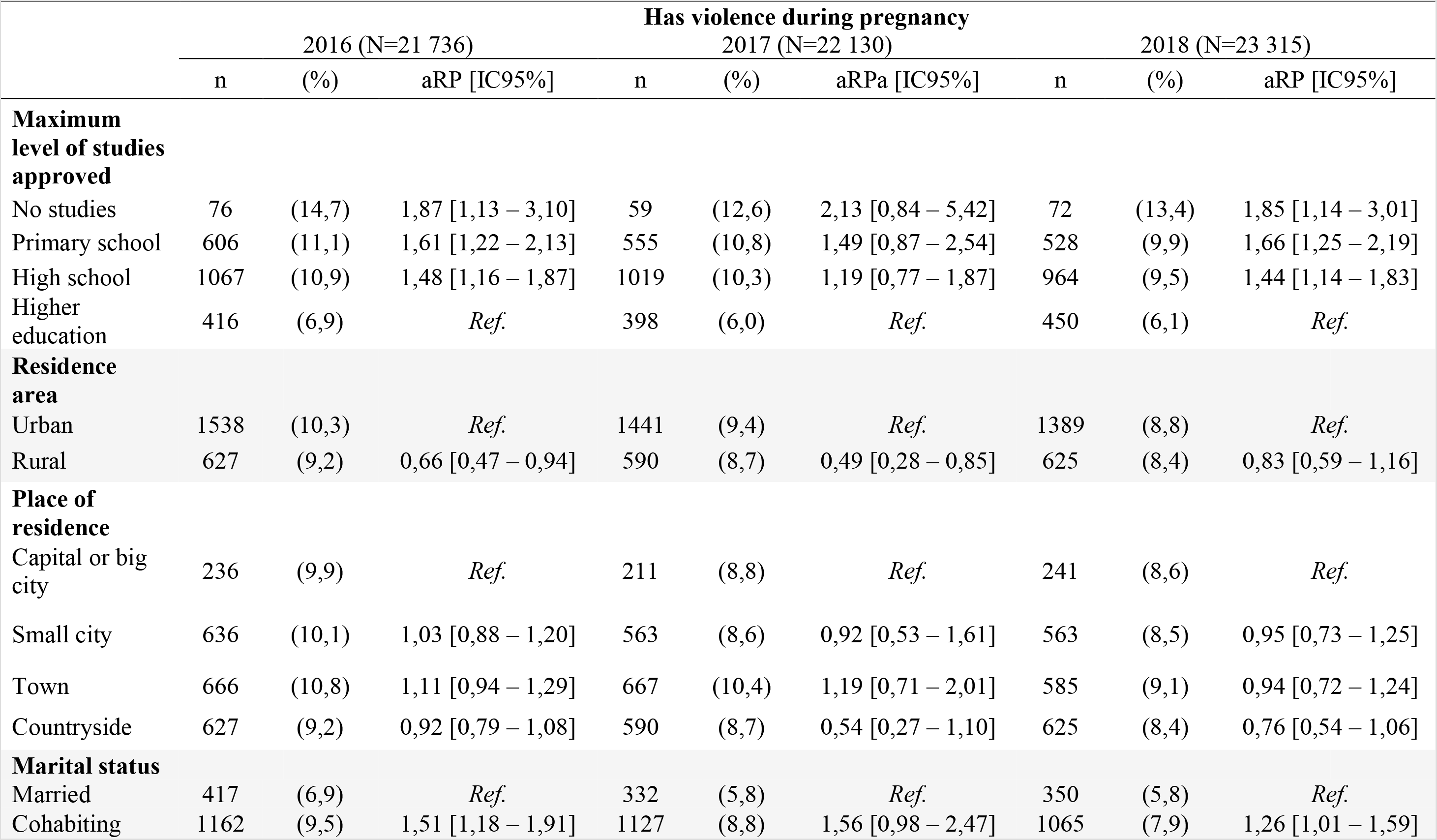
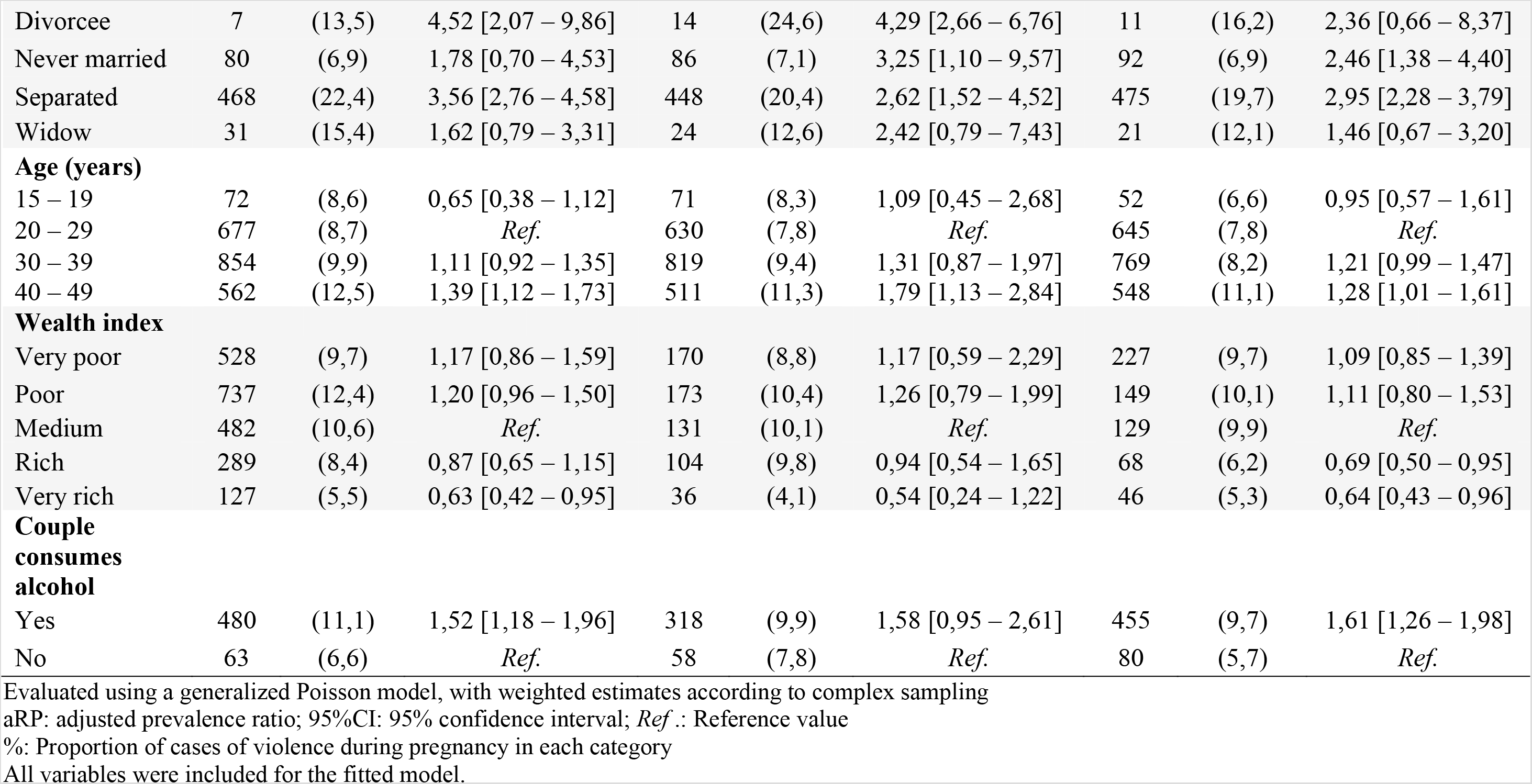
Sociodemographic factors associated with violence during pregnancy in Peru, 2016 – 2018.

Finally, the proportion of violence during pregnancy was homogeneously reduced in a large part of the evaluated variables; except in the area of residence, where there was a greater reduction in those who live in urban settings (−1.58%) than in rural settings (−0.76%), and in the wealth index, where those who were categorized as “very poor” showed an increase in violence during this period (+0.04%).

## Discussion

The proportion of violence during pregnancy found is similar to that reported in previous studies^4^, although studies that have developed specific screening strategies during prenatal care have found values close to 30%^5,7^. In Peru, the proportion of violence during pregnancy has been decreasing, which may be due to the policies implemented during 2015 and 2016 against gender violence. These sought to modify sociocultural patterns that legitimize violence (strengthen capacities in schools, promote information in the media, empower community agents and involve new actors), guarantee access to services (generate a comprehensive process for prevention, care and protection of assaulted persons, strengthen the capacities of operators, re-educate the aggressors and establish an information system) and establish legal sanctions for those who violate women^14^.

The proportion of physical violence during pregnancy was also evaluated in each region, where it is difficult to establish a direct comparison due to the lack of information in this regard. However, the national observatory reports that the highest proportions of gender violence are found in Apurímac, Junín, Puno and Cusco, which are regions that in the present study showed a high proportion of violence during pregnancy during the 3 years evaluated^15^. It was also observed that those who come from rural areas and with an extremely poor wealth index reduced the proportion of violence in low amounts or even increased it. This is due to the fact that low-income participants are less likely to have optimal prenatal care, either because of the distance to the facility or because of the control that the couple usually exerts in environments of economic dependency, thus avoiding screening and intervention. timely^16,17^

Regarding the associated factors, education in the pregnant woman showed to be decisive in the presence of violence, where a higher academic degree reduces the probability of physical abuse, which coincides with previous national studies^18^. Likewise, presenting a marital status divorced and separated showed to be strong risk factors. In this regard, a systematic review indicates that this is due to the fact that much of the violence begins in the period of separation^19^; Therefore, due to the cross-sectional nature of the study, it is possible that the rupture of the link may have occurred after the end of the pregnancy. On the other hand, people who had a pregnancy older than 40 years had a higher proportion of violence; range in which there is a low probability of requesting help from the family environment, which increases the risk of repeated episodes of violence^7^.

Finally, the present study identifies populations susceptible to violence during pregnancy, where it is essential to strengthen screening for violence, which, although it is contemplated in the perinatal card, generates a superficial search; therefore, it is also necessary to evaluate the effectiveness of the tools used^20^. Likewise, it is recommended that the consultation on violence address previous years 6 and, if identified, strengthen work with family members^7^.

Regarding the limitations of the study, the collection of information was carried out by means of a self-report and through a single question, with which there may be an underestimation in the proportion found, this could be favoured if we consider that there may be a bias of social desirability, where the participant responds according to what is socially acceptable; likewise, the absence of temporary records of the violence events and / or the memory bias that the participants present do not allow determining causality between the variables; However, we consider that the findings give us an approximation to the situation of violence during pregnancy at the national level and allow estimating populations at risk, also evaluating how these indicators have varied in recent years.

We conclude that the proportion of violence during pregnancy has been decreasing lately and has reached 8.64% in 2018, although there are regions in the south of the country that maintain values above 10%. Among the associated factors constant over time are the wealth index, the educational level of the pregnant woman, marital status, and the age range of the pregnant woman.

## Data Availability

the data are freely available on the website of the National Institute of Statistics and Informatics (http://iinei.inei.gob.pe/microdatos/).

## Information and declarations

1. Conflict of interest: The authors declare that we have no conflicts of interest
2. Ethical approval: Because it is a secondary-based analysis, approval by an ethics
committee was not required.
3. Funding sources: This research was self-financed

## Notes

### Competing Interest Statement

The authors have declared no competing interest.

### Author Declarations

Document reviewed and approved by the ethics committee of the Universidad Nacional Mayor de San Marcos.

